# Monkeypox treatment with tecovirimat in the Central African Republic under an Expanded Access Programme

**DOI:** 10.1101/2022.08.24.22279177

**Authors:** Festus Mbrenga, Emmanuel Nakouné, Christian Malaka, Josephine Bourner, Jake Dunning, Guy Vernet, Peter Horby, Piero Olliaro

## Abstract

**Background:** There is currently no specific treatment recommended for monkeypox. This expanded access programme (EAP) aims to provide tecovirimat to patients with monkeypox and collect data on patient treatment, disease evolution and outcomes under a protocol to contribute to the evidence-base for the use of the drug for monkeypox.

**Methods:** Patients with confirmed monkeypox received tecovirimat according to the recommended dosing. Data on clinical signs and symptoms were recorded daily during treatment and at follow-up visits. Blood or lesion samples were taken during treatment and at day 28 to assess viral presence of monkeypox by PCR. As tecovirimat is administered via an EAP, outcome measures were not predefined. Adverse events and clinical outcomes were monitored by evaluating the total number and location of lesions, temperature, degree of incapacity, presence of adverse events, patient survival, and viral presence throughout treatment and follow-up.

**Results:** We report outcomes in 14 patients who were enrolled between December 2021 and February 2022. Muscle pain, headache, lymphadenopathy, lesions, fever, back pain, and upper respiratory symptoms were commonly reported at admission and during follow-up. The rate of appearance of active lesions gradually decreased throughout treatment, with the median time to no new lesions being 5 days following the start of treatment. No death attributable to monkeypox occurred in this cohort.

**Conclusions:** Data collected through this EAP can help improve our knowledge about the use of tecovirimat for monkeypox. We have been able to document systematically the presentation and clinical and virological evolution of monkeypox under treatment.

**Registration number:** ISRCTN43307947

## Introduction

Monkeypox is a viral zoonotic disease caused by monkeypox virus (MPXV), a species of the Orthopoxvirus genus, which includes three other species that infect humans: cowpox virus, vaccinia virus and variola virus.^1^ Two geographically distinct clades of the MPXV have been identified, the West African clade and the Congo Basin clade, with case fatality ratios of ∼4% and 11%, respectively.^2,3^A new lineage has also recently been identified in Europe during the 2022 outbreak.^4^ Death occurs most frequently in children, pregnant women and individuals with underlying conditions. ^5,6^ Many mammals are known to carry MPXV, particularly species of rodents, although a definitive natural reservoir is yet to be identified. Transmission to humans typically occurs following contact with an infected animal or contaminated surfaces,^7^ with further onward human-to-human transmission through close contact.^8-12^

Cases of monkeypox most commonly occur in rural areas of West and Central Africa ^13,14^, but exported cases are occasionally reported in other regions,^15-17^ and concerns have been raised about the disease spreading to wider areas.^10^ Indeed, since 7 May 2022, a multi-country outbreak of West African clade monkeypox has been reported for 28 previously non-endemic countries, with evidence of community transmission and more cases likely to be reported.^18^

Cases of monkeypox typically present with a febrile prodrome with onset of a characteristic vesiculopustular rash, lymphadenopathy, and other non-specific symptoms. ^8,19,20^ However, disease presentation varies between clade; the Congo Basin clade has been associated with a pronounced rash and severe outcomes, whereas the West African clade is typically associated with milder symptoms.^21^ The emerging lineage of the 2022 outbreak has been characterised by more localised lesions – commonly reported on the genital or perianal regions of the body and associated with pain, sore throat and oedema – and does not exhibit the same prodromal symptoms as the other clades.^22,23^ However, cases falling under medical observation are likely to represent a small proportion of all monkeypox cases, many and most of which occur mostly outside the reach of affected countries’ healthcare systems.

Currently there is no specific treatment recommended for monkeypox. Cases in endemic countries are managed with supportive care to alleviate disease symptoms. ^24^ Potential antiviral compounds exist, however, and have undergone early phase testing, although no compound has yet been evaluated in a large-scale controlled clinical trials.^25^

Tecovirimat (TPOXX or ST-246^®^, SIGA Technologies, Inc., New York, NY, USA) has been identified as a potential candidate for the treatment of monkeypox.^26^ It is a small-molecule antiviral drug and was originally developed and subsequently approved in accordance with the Food and Drug Administration (FDA) Animal Efficacy Rule for the treatment of smallpox.^27,28^ Tecovirimat is now also approved by the European Medicine Agency (EMA)^29^ for the treatment of orthopoxvirus disease (smallpox, monkeypox, cowpox) and for vaccinia complications in adults and children weighing at least 13 kilograms.

Sixteen clinical studies, from Phase I through to Phase IV, have been conducted to evaluate the safety and pharmacokinetics of tecovirimat in healthy adult subjects and demonstrated good general tolerability. Tecovirimat has also been provided for emergency use under U.S. Emergency Investigational New Drug (EIND) applications in 7 cases, as well as 1 compassionate use case in Europe and 6 named patient cases (5 cases in Europe and 1 case in India).^30^ Two of these cases were in patients with confirmed monkeypox virus infection; the remainder were in patients with cowpox, neurodermitis, accidental exposure to vaccinia virus, suspected vaccinia, keratoconjunctivitis and eczema vaccinatum.^30^ However, no adequate and well-controlled studies involving patients with monkeypox, pregnant women or children have yet been conducted.

In this article, we present the results of the first phase of an Expanded Access Programme (EAP) of tecovirimat for monkeypox conducted in the Central African Republic (CAR) from December 2021 to February 2022. The study was set up in the Lobaye district of CAR, where cases typically peak mostly around the ‘caterpillar season’ between July and December, when people are more exposed to wildlife. Cases tend to occur in small clusters of <10 cases within households or communities, indicating human-to-human transmission. Overall, approximately 100 cases have been reported across the country since 2000 ^11^, although the true incidence of monkeypox in CAR is likely much higher.

This EAP aims to provide tecovirimat to patients with monkeypox and collect data on treatment, disease evolution and outcomes under a study protocol to contribute to the evidence base for the use of the drug for monkeypox. While consideration was given to a clinical trial of tecovirimat in 2020, it was considered to be infeasible at the time, due to low patient numbers and challenging field conditions in CAR. The current EAP was therefore set up with the additional aims to strengthen clinical research capacity in CAR and better understand disease incidence and presentations, in view of a possible, future randomised controlled trial, should the conditions to conduct a trial change and findings from the EAP be favourable.

The EAP is established under a tripartite collaboration agreement signed by the CAR Ministry of Health and population, Institut Pasteur Bangui (IPB), and the University of Oxford, which acts as the sponsor.

This is the first report of monkeypox cases treated systematically under a research protocol with tecovirimat in a monkeypox-endemic country.

## Methods

The EAP is underway in the Lobaye district of CAR, located in the south-west of the country bordering the Republic of Congo and the Democratic Republic of the Congo. Mbaïki hospital – the district primary and referral health care facility with good access to the capital city Bangui – was selected as the main treatment centre at which tecovirimat would be administered for the EAP. Communication and travel within this highly rural, sparsely populated, and densely forested region is challenging, which has implications both for case finding and referral. While cases are reported also from other parts of CAR, the security situation in the country is such that the Lobaye district was the only practicable option.

A suspected case is defined as a subject presenting with clinical signs and symptoms indicative of monkeypox, such as fever and characteristic rash. We set in place an intensified active surveillance system: suspected cases are identified through referrals from community health workers, other health facilities and community leaders, and through contact tracing in villages of the district by the EAP team. Following the identification of a suspected case, blood and lesion samples are taken from the subject and blood samples are also taken from close contacts (regardless of the presence of clinical signs and symptoms) and transferred to the Institut Pasteur de Bangui reference laboratory for case confirmation. Eligible patients are those who received laboratory confirmation of monkeypox infection and weighed ≥13 kg. Patients receiving repaglinide are ineligible due to risk of interactions with tecovirimat.

Following consent, adult patients receive 600 mg of oral tecovirimat (3 × 200 mg capsules) twice daily for 14 days. Dosing for paediatric patients is based on weight: children weighing 13kg to <25kg receive 200 mg of tecovirimat (1 capsule) twice daily for 14 days; children weighing 25kg to <40 kg receive 400 mg of tecovirimat (2 capsules) twice daily for 14 days; and children weighing ≥40 kg receive 600 mg of tecovirimat (three capsules) of tecovirimat twice daily for 14 days. Patients are to stay in hospital for the duration of treatment, with follow-up visits planned on days 15 and 28. Data on clinical signs and symptoms, including lesion burden, are recorded daily during treatment and at each follow-up visit.

Blood or lesions samples on pus or crusts are scheduled on days 1, 4, 8 and 14 during treatment, and then at day 28, to assess viral presence of MPXV using the G2R-G real-time PCR assay and the Congo Basin clade of the virus using the C3L real-time PCR assay.^31^ In this cohort, multiple samples were taken from some patients at some study visits. Patients with positive PCR on day 14 had an additional sample on day 21. Monkeypox disease is confirmed by detecting viral DNA on blood samples and/or lesion swabs.

As tecovirimat is administered via an EAP, outcome measures and endpoints were not predefined. Adverse events and clinical outcomes are monitored by evaluating the total number and location of lesions, temperature, degree of incapacity, presence of adverse events, patient survival, and virus DNA levels throughout treatment and follow-up.

The EAP was approved by the Ministry of Health, Central African Republic, and obtained ethical clearance by the national ethics committee (“Comité Ethique et Scientifique, Université de Bangui Faculté des Sciences de la Santé”) and Oxford Tropical Research Ethics Committee (OxTREC) at the University of Oxford.

The EAP is registered on the ISRCTN registry (reference: ISRCTN43307947).^32^

## Statistical analysis

A descriptive analysis of the demographics, signs and symptoms, and patient outcome of the enrolled patients who were treated with tecovirimat is presented. Signs and symptoms are reported as the number and proportion of patients for whom each sign or symptom was recorded at admission (baseline) and at any time post-baseline. A denominator is provided for each variable to indicate the number of patients for whom data were available.

Lesion presence is summarised as the number and proportion of patients for whom lesions are reported overall and by location at admission and any time post-baseline. At point of data collection, lesions were categorised as either active lesions, scabs or scars and are summarised here as either the presence of active lesions or the presence of lesions of any type. Lesion burden is summarised on a categorical scale: None; 1-5 lesions; 6-25 lesions; 26-100 lesions; >100 lesions. The median time to no active lesions is also reported.

The number and proportion of patients testing positive on PCR for MPXV, G2R-G and the Congo Basin clade, is reported for all study timepoints overall and per sample type tested (blood, active lesion and crust). The mean, standard deviation and range of reported CT values are also presented where more than one sample is available at any given timepoint.

A summary of serious adverse events (SAEs), including number of SAEs per patient and severity, is also provided.

The analysis was conducted by JB.

## Results

Between December 2021 and February 2022, 14 patients with laboratory-confirmed monkeypox were enrolled and treated with tecovirimat. Of the patients enrolled, four were male and ten were female with a median age of 23 years (range 4 to 38 years) (**Table 1**). None of the female patients were known to be pregnant at the point of enrolment. Three (21%) of the enrolled patients were under the age of 16.

**Table 1.** Patients’ demographics and characteristics at baseline and post-baseline.

|  | Baseline | Day 4 | Day 8 | Day 14 | Day 28 |
| --- | --- | --- | --- | --- | --- |
| ***Demographic characteristics*** |  |  |  |  |  |
| Male : female | 4 : 10 |  |  |  |  |
| Age (years): median (range) | 23 (4 - 38) |  |  |  |  |
| ***Comorbidities*** |  |  |  |  |  |
| Malaria: n/N (%) patients | 11/13 (85%) |  |  |  |  |
| HIV: n/N (%) patients | 1/3 (7%) |  |  |  |  |
| ***General characteristics*** |  |  |  |  |  |
| Time (days) from symptom onset to treatment start: median (range) | 21 (5-45) |  |  |  |  |
| ***Signs and symptoms*** | **n/N (%) patients** |  |  |  |  |
| Fever (temperature >38.0 °C) | 9/14 (64%) | 4/14 (29%) | 0/14 (0%) | 0/14 (0%) | 1/13 (8%) |
| Lesions: | 14/14 (100%) | 14/14 (100%) | 14/14 (100%) | 14/14 (100%) | 13/13 (100%) |
| New lesions: | 10/14 (71%) | 11/14 (79%) | 6/14 (43%) | 1/14 (7%) | 0/13 (0%) |
| Number of lesions (total): median (range) | 302 (54-7586) |  |  |  |  |
| 0-49 | 0/14 (0%) | 1/14 (7%) | 0/14 (0%) | 0/14 (0%) | 0/14 (0%) |
| 50-99 | 3/14 (21%) | 0/14 (0%) | 0/14 (0%) | 0/14 (0%) | 0/14 (0%) |
| 100-199 | 0/14 (0%) | 2/14 (14%) | 1/14 (7%) | 1/14 (7%) | 6/13 (46%) |
| 200-299 | 4/14 (29%) | 3/14 (21%) | 5/14 (36%) | 5/14 (36%) | 4/13 (31%) |
| 300-399 | 3/14 (21%) | 2/14 (14%) | 2/14 (14%) | 2/14 (14%) | 2/13 (15%) |
| 400-499 | 1/14 (7%) | 3/14 (21%) | 3/14 (21%) | 3/14 (21%) | 0/14 (0%) |
| 500-599 | 2/14 (14%) | 1/14 (7%) | 2/14 (14%) | 2/14 (14%) | 0/14 (0%) |
| >600 | 1/14 (7%) | 2/14 (14%) | 1/14 (7%) | 1/14 (7%) | 1/13 (8%) |
| Location of lesions: |  |  |  |  |  |
| Hands | 14/14 (100%) | 14/14 (100%) | 14/14 (100%) | 14/14 (100%) | 13/13 (100%) |
| Feet | 14/14 (100%) | 14/14 (100%) | 14/14 (100%) | 14/14 (100%) | 13/13 (100%) |
| Face | 14/14 (100%) | 14/14 (100%) | 14/14 (100%) | 14/14 (100%) | 13/13 (100%) |
| Back | 14/14 (100%) | 14/14 (100%) | 14/14 (100%) | 14/14 (100%) | 13/13 (100%) |
| Thighs | 14/14 (100%) | 14/14 (100%) | 14/14 (100%) | 14/14 (100%) | 13/13 (100%) |
| Legs | 14/14 (100%) | 14/14 (100%) | 14/14 (100%) | 14/14 (100%) | 13/13 (100%) |
| Arms | 14/14 (100%) | 14/14 (100%) | 14/14 (100%) | 14/14 (100%) | 13/13 (100%) |
| Forearms | 14/14 (100%) | 14/14 (100%) | 14/14 (100%) | 14/14 (100%) | 13/13 (100%) |
| Abdomen | 14/14 (100%) | 14/14 (100%) | 14/14 (100%) | 14/14 (100%) | 13/13 (100%) |
| Chest | 14/14 (100%) | 14/14 (100%) | 14/14 (100%) | 14/14 (100%) | 13/13 (100%) |
| Genitals | 11/14 (79%) | 12/14 (86%) | 12/14 (86%) | 13/14 (93%) | 12/13 (92%) |
| Mouth | 10/14 (71%) | 9/14 (64%) | 10/14 (71%) | 10/14 (71%) | 9/13 (69%) |
| Nose | 11/14 (79%) | 11/14 (79%) | 8/14 (57%) | 9/14 (64%) | 8/13 (62%) |
| Other | 13/14 (93%) | 13/14 (93%) | 14/14 (100%) | 13/14 (93%) | 11/13 (85%) |
| Lesion pain | 11/14 (79%) | 10/14 (71%) | 3/14 (21%) | 0/14 (0%) | 0/13 (0%) |
| Lesion pain score: median (range) | 7 (5-9) | 5 (2-8) | 2 (1-5) | N/A | N/A |
| Lymphadenopathy | 14/14 (100%) | 14/14 (100%) | 14/14 (100%) | 13/14 (93%) | 6/13 (46%) |
| Keratitis | 2/14 (14%) | 2/14 (14%) | 2/14 (14%) | 1/14 (7%) | 2/13 (15%) |
| Cough | 5/14 (36%) | 2/14 (14%) | 2/14 (14%) | 0/14 (0%) | 0/13 (0%) |
| Clear sputum | 3/13 (23%) | 0/14 (0%) | 2/14 (14%) | 0/14 (0%) | 0/13 (0%) |
| Upper respiratory symptoms | 11/14 (79%) | 6/14 (43%) | 1/14 (7%) | 1/14 (7%) | 0/13 (0%) |
| Lower respiratory symptoms | 4/14 (29%) | 3/14 (21%) | 2/14 (14%) | 0/14 (0%) | 0/13 (0%) |
| Breathlessness | 0/13 (0%) | 0/14 (0%) | 0/14 (0%) | 0/14 (0%) | 0/13 (0%) |
| Vomiting | 0/14 (0%) | 0/14 (0%) | 0/14 (0%) | 0/14 (0%) | 0/13 (0%) |
| Diarrhoea | 0/14 (0%) | 0/14 (0%) | 0/14 (0%) | 0/14 (0%) | 0/13 (0%) |
| Headache | 14/14 (100%) | 11/14 (79%) | 7/14 (50%) | 0/14 (0%) | 0/13 (0%) |
| Back pain | 11/14 (79%) | 13/14 (93%) | 7/14 (50%) | 0/14 (0%) | 0/13 (0%) |
| Muscle pain | 14/14 (100%) | 10/14 (71%) | 4/14 (29%) | 0/14 (0%) | 0/13 (0%) |
| Seizure | 0/14 (0%) | 0/14 (0%) | 0/14 (0%) | 0/14 (0%) | 0/13 (0%) |
| Patient is able to eat independently | 13/14 (93%) | 13/14 (93%) | 14/14 (100%) | 14/14 (100%) | 13/13 (100%) |
| Patient is able to drink independently | 14/14 (100%) | 14/14 (100%) | 14/14 (100%) | 14/14 (100%) | 13/13 (100%) |
| Patient is able to walk independently | 13/14 (93%) | 13/14 (93%) | 13/14 (93%) | 14/14 (100%) | 13/13 (100%) |
| **Vital signs** |  |  |  |  |  |
| Respiratory rate >25 | 2/14 (14%) | 1/14 (7%) | 0/14 (0%) | 0/14 (0%) | 0/13 (0%) |
| Pulse >100 | 2/14 (14%) | 2/14 (14%) | 1/14 (7%) | 0/14 (0%) | 0/13 (0%) |
| Systolic blood pressure <90mm Hg | 0/14 (0%) | 0/14 (0%) | 0/14 (0%) | 0/14 (0%) | 0/13 (0%) |

|  | **Baseline** | **Day 4** | **Day 8** | **Day 14** | **Day 28** |
| --- | --- | --- | --- | --- | --- |
| Oxygen saturation <95% | 4/14 (29%) | 3/14 (21%) | 0/14 (0%) | 0/14 (0%) | 0/13 (0%) |
| Level of consciousness |  |  |  |  |  |
| Alert | 14/14 (100%) | 11/14 (79%) | 11/14 (79%) | 11/14 (79%) | 13/13 (100%) |
| Confusion | 0/14 (0%) | 0/14 (0%) | 0/14 (0%) | 0/14 (0%) | 0/13 (0%) |
| Responsive to voice | 0/14 (0%) | 3/14 (21%) | 3/14 (21%) | 3/14 (21%) | 0/13 (0%) |
| Responsive to pain | 0/14 (0%) | 0/14 (0%) | 0/14 (0%) | 0/14 (0%) | 0/13 (0%) |
| Unconscious | 0/14 (0%) | 0/14 (0%) | 0/14 (0%) | 0/14 (0%) | 0/13 (0%) |

<sup>a</sup> Post-baseline includes any timepoint following baseline, including days on which patients are receiving treatment

At the point of admission, 11 patients tested positive for malaria on RDT and one patient subsequently tested positive for HIV.

The median time from symptom onset to start of treatment was 21 days (range 5 to 45 days).

### Signs and symptoms

At admission, all 14 (100%) patients presented with muscle pain, headache, lymphadenopathy and lesions of any type characteristic of monkeypox infection (**Fig 1, Fig S1**). Fever was reported in nine (64%) patients, and back pain and upper respiratory symptoms were reported in 11 (79%) patients. Cough, lower respiratory symptoms, clear sputum was reported and keratitis were reported less frequently, in five (36%), four (29%), three (21%) and two (14%) patients respectively.

**Fig 1.**
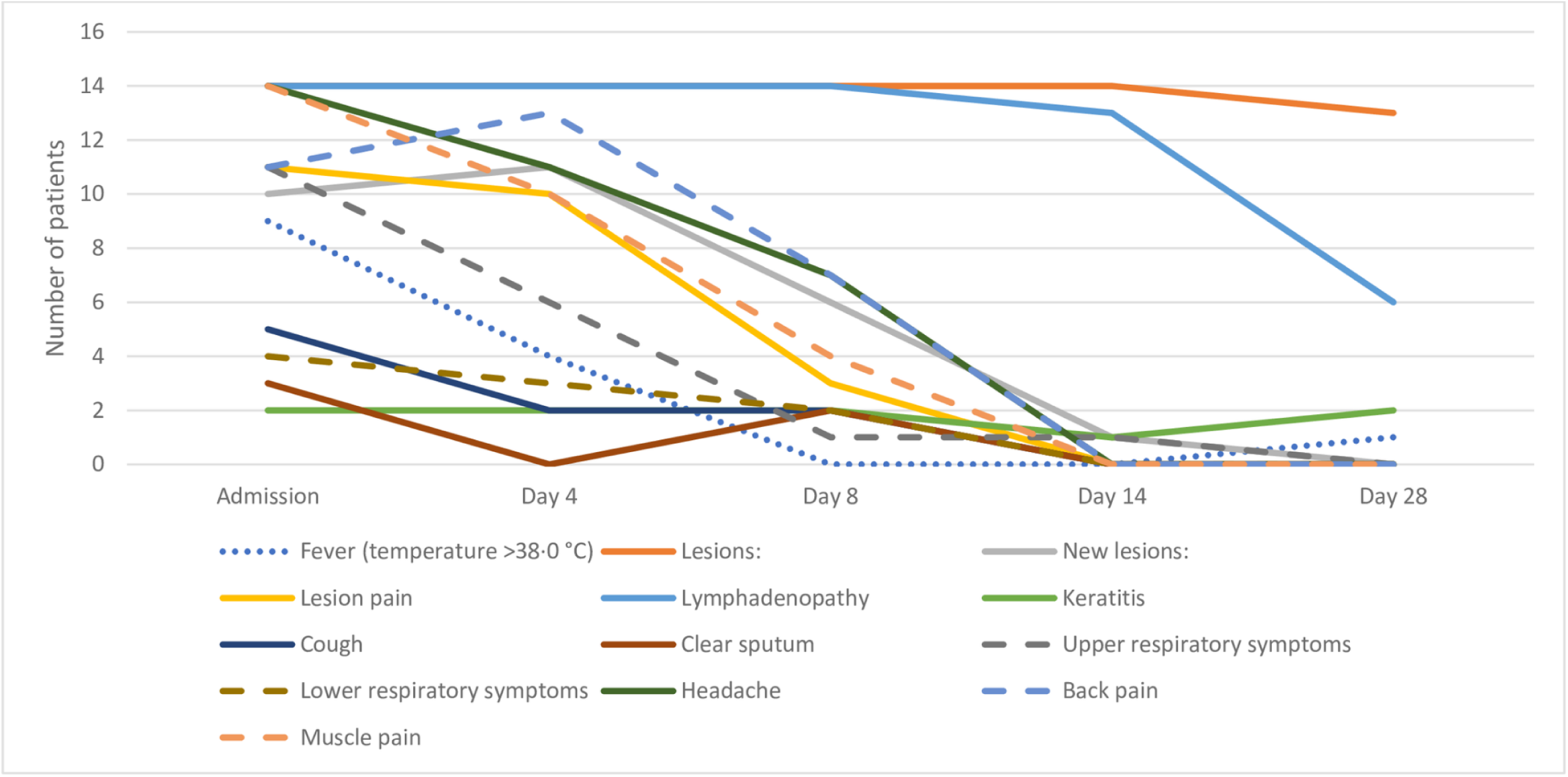
Number of patients reporting symptoms at each study timepoint.

By D4, all (100%) patients reported lymphadenopathy and lesions (any type) and more than half of enrolled patients presented with back pain (13(93%)), headache (11(79%)), new lesions (11(79%)), active lesions (10(71%) and muscle pain (10(71%)).

At D8, all (100%) patients continued to present with lymphadenopathy and lesions (any type). The numbers of symptoms experienced by more than half of all patients decreased to headache and back pain, which were each reported by 7 (50%) patients.

Most symptoms had disappeared for all patients at D14 and D28. However, lesions and lymphadenopathy persisted in the highest numbers of patients at both timepoints. Two (15%) patients had keratitis and 1 (8%) patient had a fever at D28.

An increase was seen from baseline in the number of patients reporting back pain (13 (93%)), fever (12 (86%)), active lesions (11(79%)), cough (6(43%)), and lower respiratory symptoms (5(36%)). Upper respiratory symptoms, clear sputum and keratitis were reported in the same number of patients post-baseline as at baseline. Breathlessness, vomiting and seizure were not noted at baseline, but were each reported for one (7%) patient post-baseline. Data were collected for the presence diarrhoea, but this symptom was not detected in any of the enrolled patients from admission to the final study visit (**Table 1**).

### Vital signs

On admission, four (29%) patients had oxygen saturation <95% and two (14%) patients had a respiratory rate of >25 breaths per minute and pulse >100 bpm (**Table 1**).

Post-baseline, the frequency of patients with oxygen saturation <95%, respiratory rate >25 breaths per minute and pulse >100 increased, and were recorded for 11 (79%), four (29%) and nine (64%) patients respectively.

Blood pressure measurements were taken at admission, throughout treatment and during follow-up; systolic pressure measurements below 90 mmHg were not recorded at any timepoint for any of the enrolled patients.

### Lesions

All patients presented with lesions on admission, of whom 10 (71%) presented with active lesions (**Table 1**). The number of patients presenting with active lesions decreased following start of treatment (Day 1), as did the total number of active lesions (**Fig 2**). The median time to no new lesions was five days (range 2-28 days) (**Table 2**).

**Fig 2.**
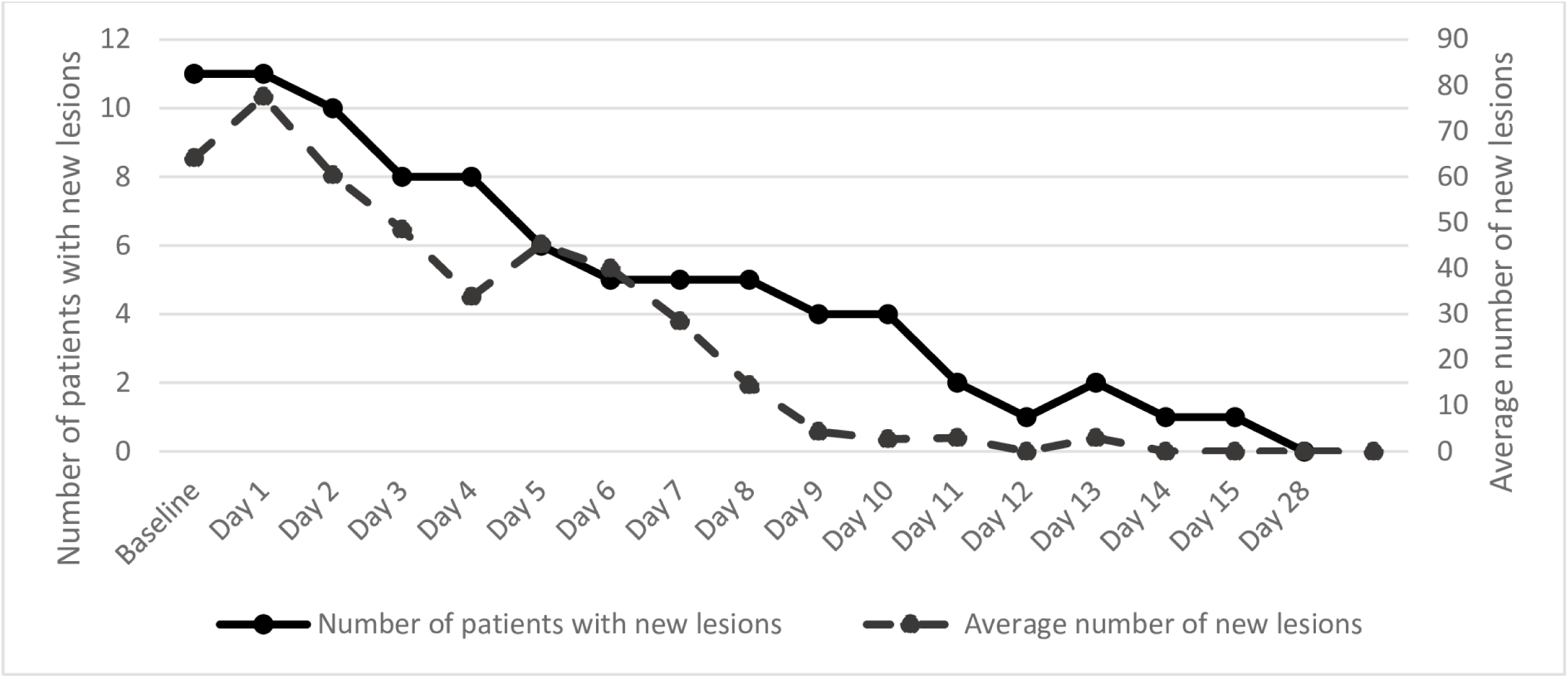
Number of patients with new lesions and average number of new lesions from baseline to day 28.

**Table 2.**
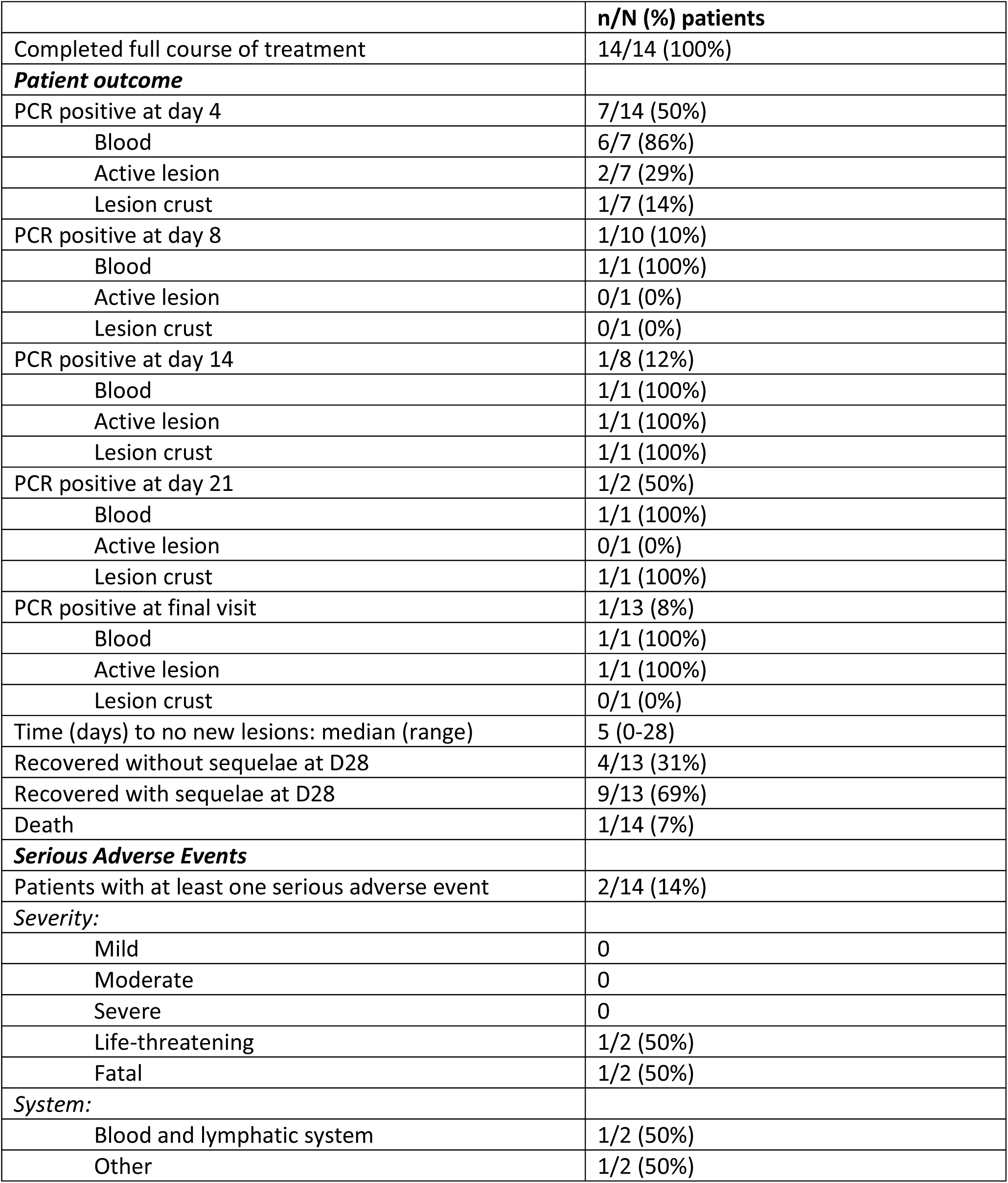
Outcomes.

|  | **n/N (%) patients** |
| --- | --- |
| Completed full course of treatment | 14/14 (100%) |
| ***Patient outcome*** |  |
| PCR positive at day 4 | 7/14 (50%) |
| Blood | 6/7 (86%) |
| Active lesion | 2/7 (29%) |
| Lesion crust | 1/7 (14%) |
| PCR positive at day 8 | 1/10 (10%) |
| Blood | 1/1 (100%) |
| Active lesion | 0/1 (0%) |
| Lesion crust | 0/1 (0%) |
| PCR positive at day 14 | 1/8 (12%) |
| Blood | 1/1 (100%) |
| Active lesion | 1/1 (100%) |
| Lesion crust | 1/1 (100%) |
| PCR positive at day 21 | 1/2 (50%) |
| Blood | 1/1 (100%) |
| Active lesion | 0/1 (0%) |
| Lesion crust | 1/1 (100%) |
| PCR positive at final visit | 1/13 (8%) |
| Blood | 1/1 (100%) |
| Active lesion | 1/1 (100%) |
| Lesion crust | 0/1 (0%) |
| Time (days) to no new lesions: median (range) | 5 (0-28) |
| Recovered without sequelae at D28 | 4/13 (31%) |
| Recovered with sequelae at D28 | 9/13 (69%) |
| Death | 1/14 (7%) |
| ***Serious Adverse Events*** |  |
| Patients with at least one serious adverse event | 2/14 (14%) |
| ***Severity:*** |  |
| Mild | 0 |
| Moderate | 0 |
| Severe | 0 |
| Life-threatening | 1/2 (50%) |
| Fatal | 1/2 (50%) |
| ***System:*** |  |
| Blood and lymphatic system | 1/2 (50%) |
| Other | 1/2 (50%) |

At admission, throughout treatment and follow-up, lesions were reported on the hands, feet, face, back, thighs, legs, arms, forearms, abdomen and chest for all patients (**Table 1**). Eleven patients (79%) had lesions on the genitals or nose, increasing to 12 (86%) patients post-baseline, and 10 patients (71%) reported lesions on the mouth from baseline to the final visit.

The highest lesion burden was found on the legs (**Fig 3**), on which patients had a median lesion burden of 32 at admission, followed by the back and face, on which patients had a median lesion burden of 24 and 23 lesions at admission respectively. The lowest lesion burden was found on the mouth, which had a median lesion burden of zero to four lesions from admission to the final study visit.

**Fig 3.**
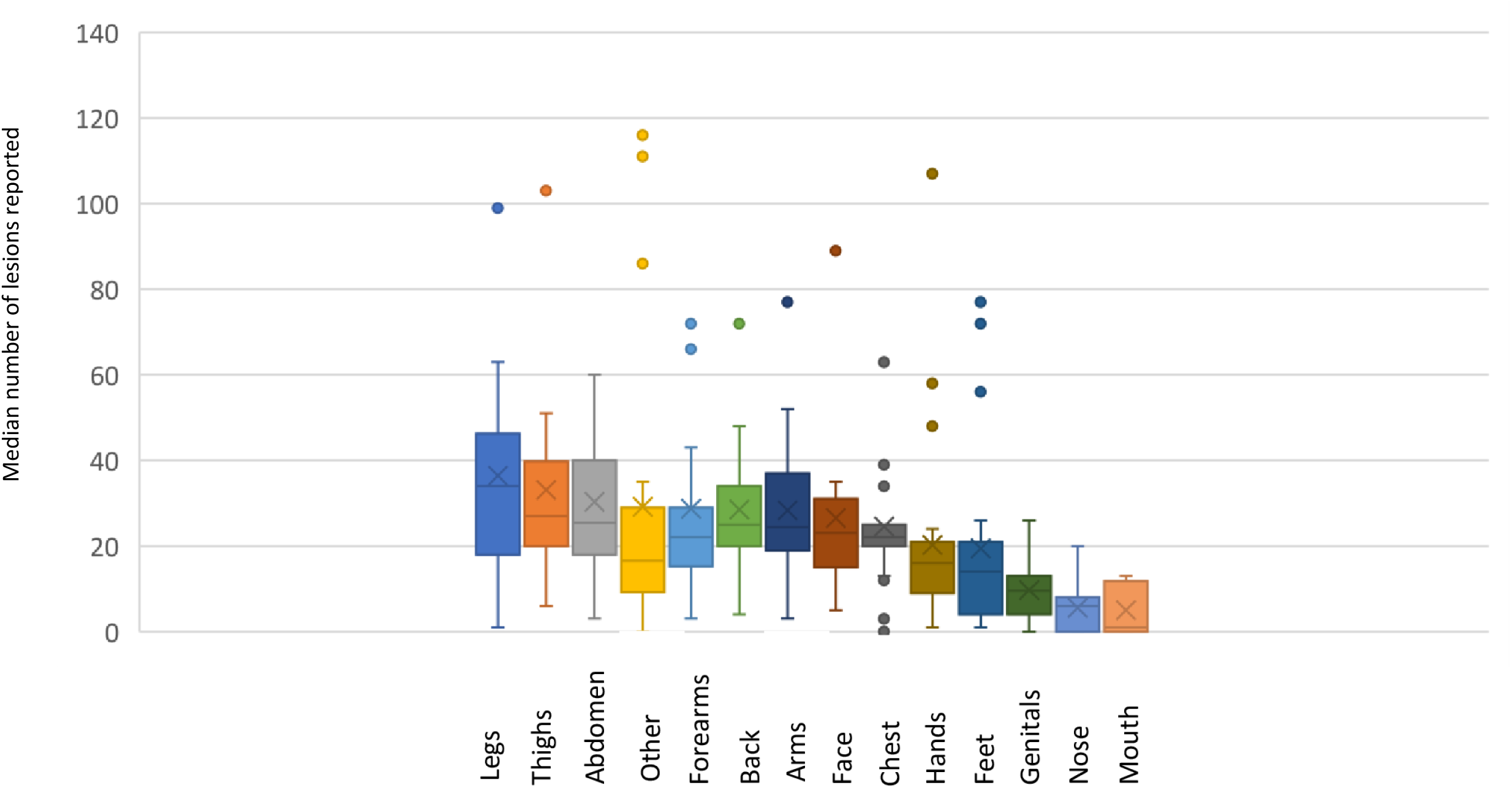
Number of lesions per area of the body.

### Virological outcomes

At baseline, all patients had a blood sample taken; six patients also had a lesion sample taken. All patients tested positive for MPXV on at least one sample (**Table 2, Table S2**) and 12 (86%) patients tested positive for the Congo Basin clade (**Table S3**).

Twelve (86%) patients tested positive on blood with a mean CT of 32 (range 21 to 42) (**Table S2**). Six (43%) patients tested positive on a lesion sample with a mean CT of 28 (range 18 to 39). On Day 4, seven (50%) patients received a positive PCR result for MPXV on at least one sample (**Table S2; Fig S4**). Six patients tested positive on blood and one patient tested positive on a lesion sample. Blood samples had a mean CT value of 38 (range 36 to 39).

From Day 8 onwards, one patient remained positive on all samples taken up to and including their final visit (**Table S2**).

Due to field challenges, it was not possible to collect samples or report CT values for all patients at each timepoint.

### Overall outcomes

All enrolled patients completed a full course of treatment with tecovirimat (**Table 2**).

Due to logistical constraints, as patients live in remote areas that are difficult to access, the D28 visit was conducted between D19 and D55.

No correlation was found between duration of illness prior to treatment start and time to lesion resolution (R^2^ = 0.21), between duration of illness and CT on blood at diagnosis (R^2^ = 0.09), and between baseline CT and time to lesion resolution (R^2^ = 0.001).

One fatality was reported among the enrolled patient population (see serious adverse events below). Four (31%) patients recovered without sequelae and nine (69%) patients recovered with sequelae. In all cases the sequelae were reported as scarring as a result of lesions.

### Serious adverse events

Two serious adverse events (SAEs) were reported during the course of the programme for two patients (**Table 2**).

One patient experienced life-threatening anaemia on Day 9 of treatment. On admission, the patient tested positive for malaria and a haematocrit test returned a reading of 20%. The patient’s haematocrit worsened to 18% on Day 9. Investigations carried out for this event also resulted in laboratory confirmation of HIV, for which antiretroviral treatment was started. The patient received a blood transfusion to treat the anaemia and recovered without sequelae on Day 13. The medical monitor for the programme considered that this event was not related to tecovirimat.

The second SAE resulted in death, which is being treated as unexplained and occurred during follow-up on Day 17. The patient had been discharged on day 14, at which time they were doing well, all lesions resolved, and tested PCR negative. The medical monitor considered that, in the absence of an official report on the cause of death, the event was unlikely to be related to tecovirimat.

## Discussion

This is the first report of protocolised, consented use of a treatment for monkeypox. Evidence supporting the use of tecovirimat has so far been extrapolated from in vitro and animal studies, due to the challenges of conducting clinical trials in the countries where the disease occurs, often in unstable areas. While the safety and tolerability of tecovirimat had been established in 973 healthy volunteers and 70 patients with either hepatic or renal impairment, before this EAP took place, only 14 patients had received tecovirimat for treatment under named patient or compassionate use – two of which were patients with confirmed monkeypox virus infection.^30,33,34^. For the first time, under this EAP patients with monkeypox are treated and followed up following a preestablished clinical protocol.

In patients enrolled in this EAP, muscle pain, headache, lymphadenopathy, lesions, fever, back pain and upper respiratory symptoms were commonly reported at admission and during follow-up. The rate of appearance of active lesions gradually decreased throughout treatment, with the median time to no new lesions being 5 days following the start of treatment.

The primary limitations of this report are the small number of cases enrolled and the late presentation of a number of the enrolled cases. An additional confounding factor is the high proportion of enrolled patients who had concomitant malaria infection. As only 14 patients were enrolled, no conclusions can be drawn about safety and it is not possible to generalise the outcomes of the study population to the wider monkeypox patient population. In CAR, treating cases early is challenging, due to a combination of factors: monkeypox occurs in remote areas, often far outside the reach of the health system; limited awareness of the disease in the communities and among healthcare workers; time-lag between a case being suspected, sample taken, and laboratory confirmation since samples must travel all the way to the reference laboratory in Bangui. For these 14 patients, it took a median time of three weeks from symptoms onset to be treated, ranging broadly from 5 to 45 days. The signs and symptoms presented here may not be representative of the earlier stages of monkeypox infection. There could therefore be a selection bias for cases who would tend to have a favourable resolution on their own. DNA levels were generally low, as derived from a mean CT value of 32 at diagnosis. However, no correlation was found between prior duration of illness and time to lesion resolution, between prior duration of illness and baseline DNA levels, and between baseline DNA levels and time to lesion resolution.

No death attributable to monkeypox occurred in this cohort while the Congo basin clade reportedly carries a risk of fatal disease (estimated at 10.6% (95% CI: 8.4%– 13.3%) vs. West African 3.6% (95% CI: 1.7%– 6.8%)).^3^

Data collected through this EAP can help improve our knowledge of the use of tecovirimat in the conditions of use in a country endemic for the Congo clade of the monkeypox virus. Setting this programme in place has also brought other significant additions. We have been able to document systematically the presentation and clinical and virological evolution of monkeypox under treatment; we have increased clinical research capacity in the country; importantly, we have successfully piloted an outreach and awareness programme and an active, community-based surveillance system to identify and channel cases for treatment that is sustainable and can be expanded.

The study team plans to continue enrolment for at least one more monkeypox transmission season in CAR.

## Data Availability

All data produced in the present study are available upon reasonable request to the authors

## Acknowledgments / Conflicts of interest and financial disclosures

The authors declare no conflict of interest. SIGA was notified of, but not involved in the design, execution, analyses and reporting on the programme. SIGA donated 100 treatments for use under the EAP. The EAP was financed with core funds of IPB, financial support to IPB from SIGA, the University of Oxford under the UK Foreign, Commonwealth and Development Office and Wellcome (215091/Z/18/Z) and the Bill & Melinda Gates Foundation (OPP1209135). For the purpose of Open Access, the author has applied a CC BY public copyright licence to any Author Accepted Manuscript version arising from this submission. The University of Oxford is the sponsor of the programme.

---

**Fig S1.**
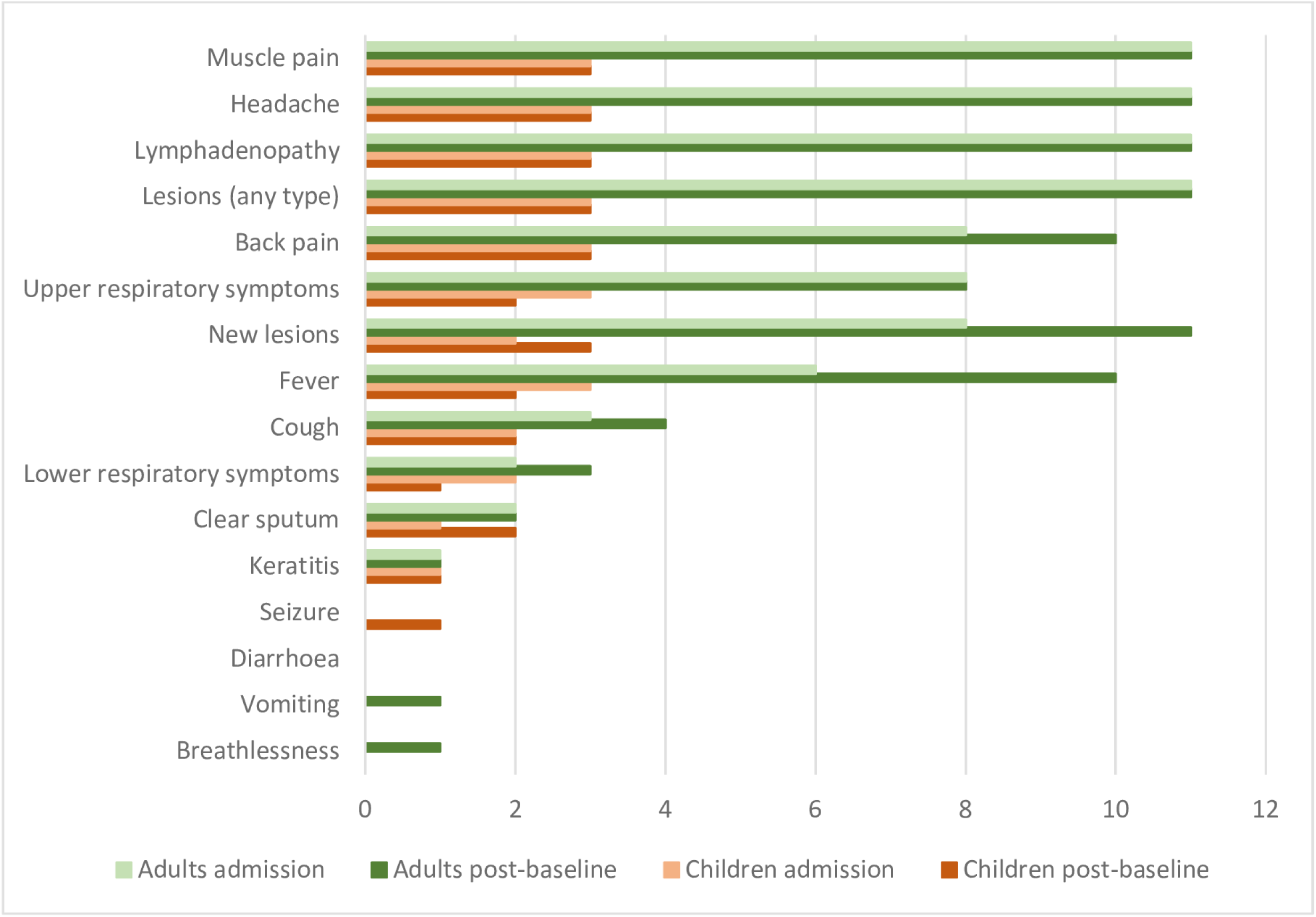
number of adult and paediatric patients reporting symptoms at baseline and any time post-baseline.

**Table S2.**
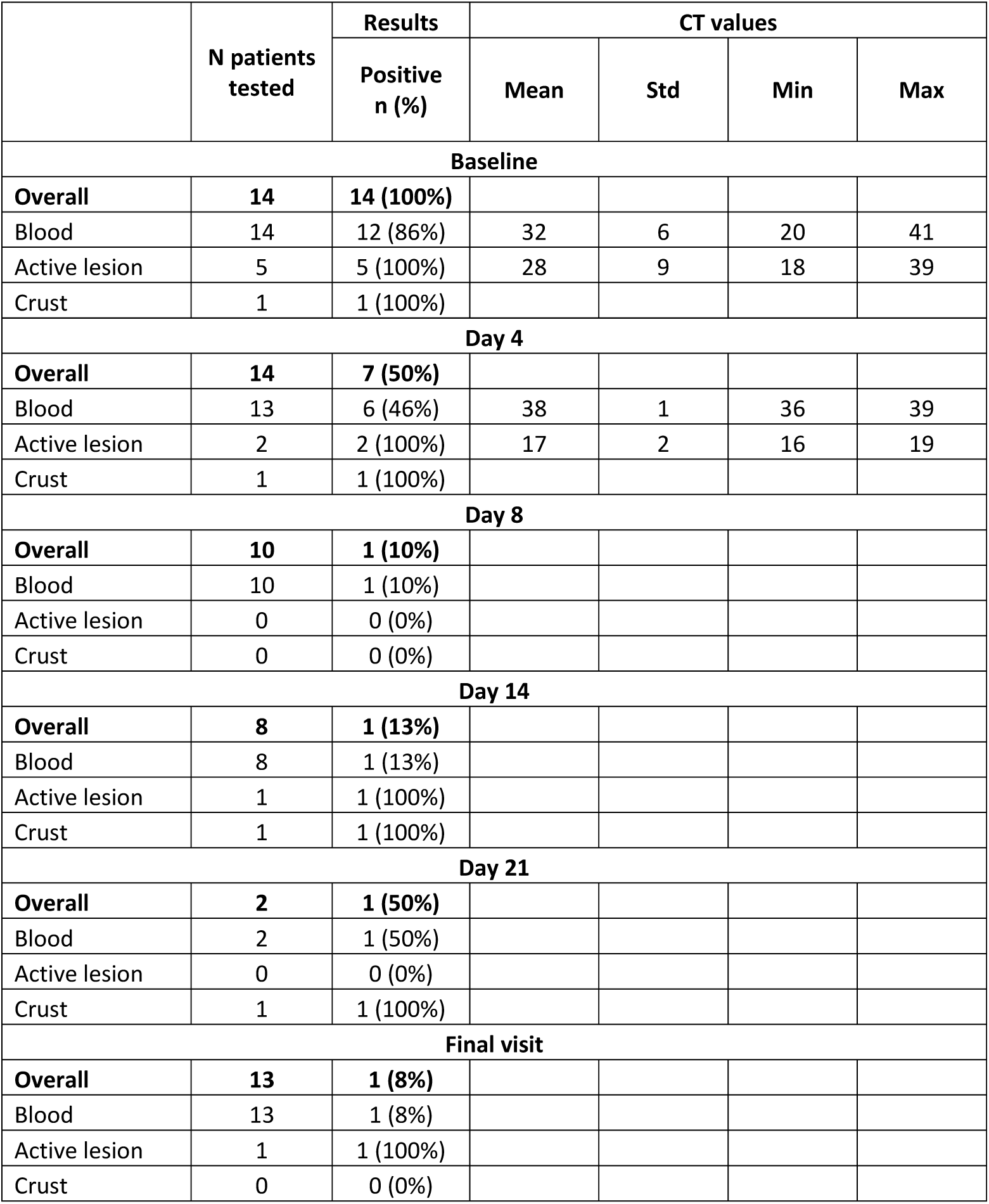
Virological outcomes using G2R-G real-time PCR assay.

**Table S3.**
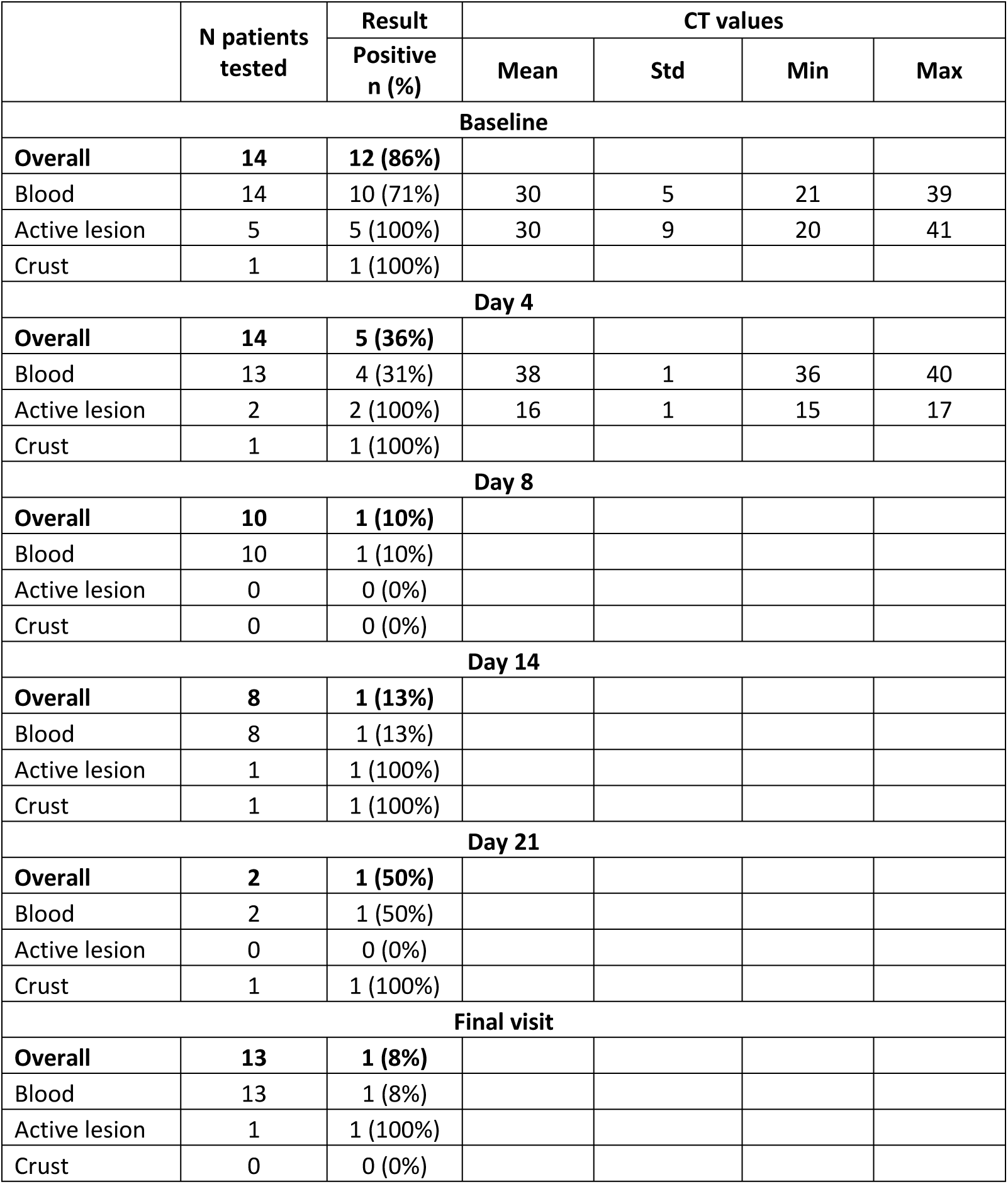
Virological outcomes using C3L real-time PCR assay.

**Fig S4.**
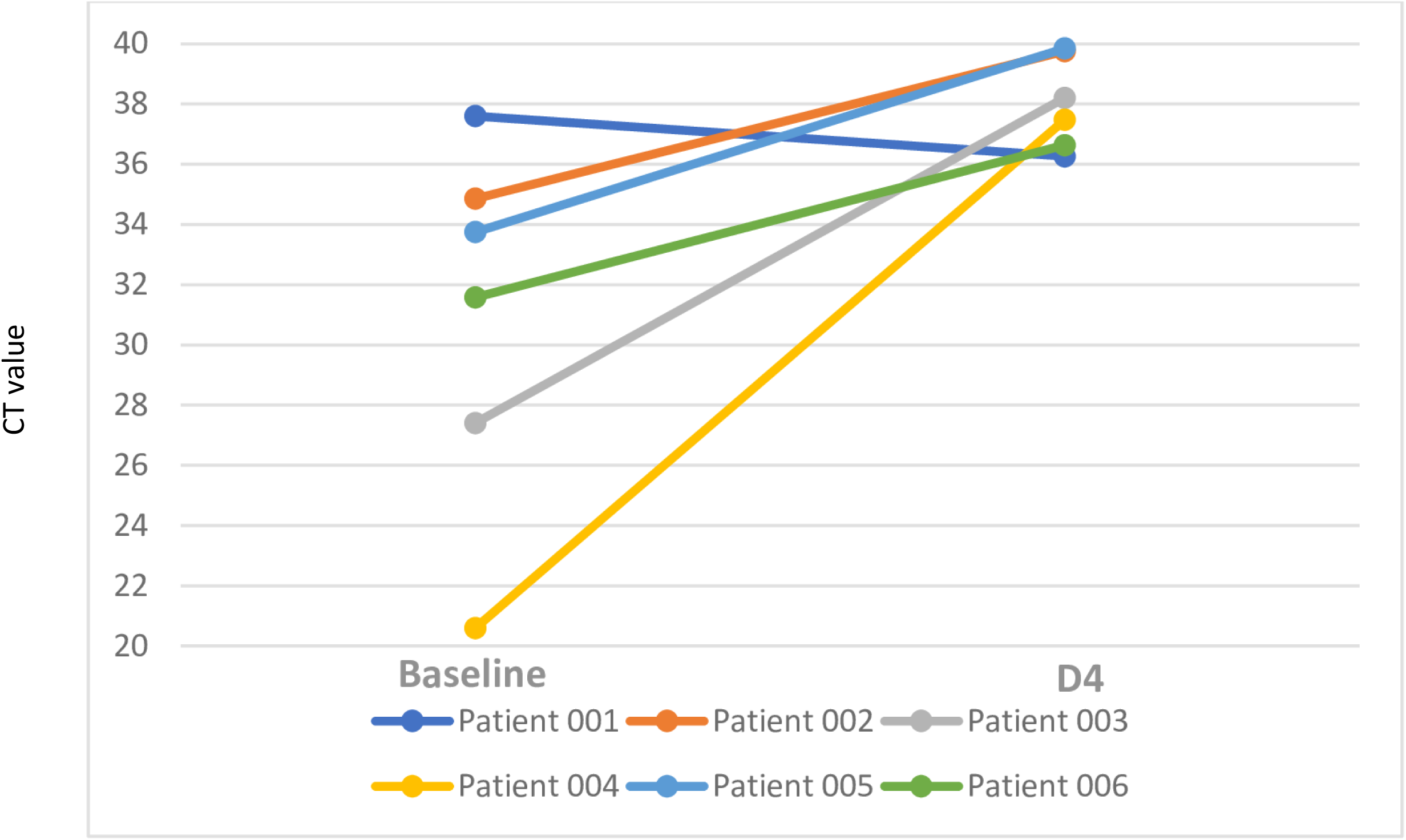
CT values of six patients from baseline to D4.

## Notes

### Competing Interest Statement

The authors have declared no competing interest.

### Clinical Trial

ISRCTN43307947

### Author Declarations

The ethics committees of the University of Oxford and University of Bangui gave ethical approval for this work.

