## Supplementary file for "Monkeypox treatment with tecovirimat in the Central African Republic under an Expanded Access Programme"

**Fig S1 – number of adult and paediatric patients reporting symptoms at baseline and any time post-baseline**

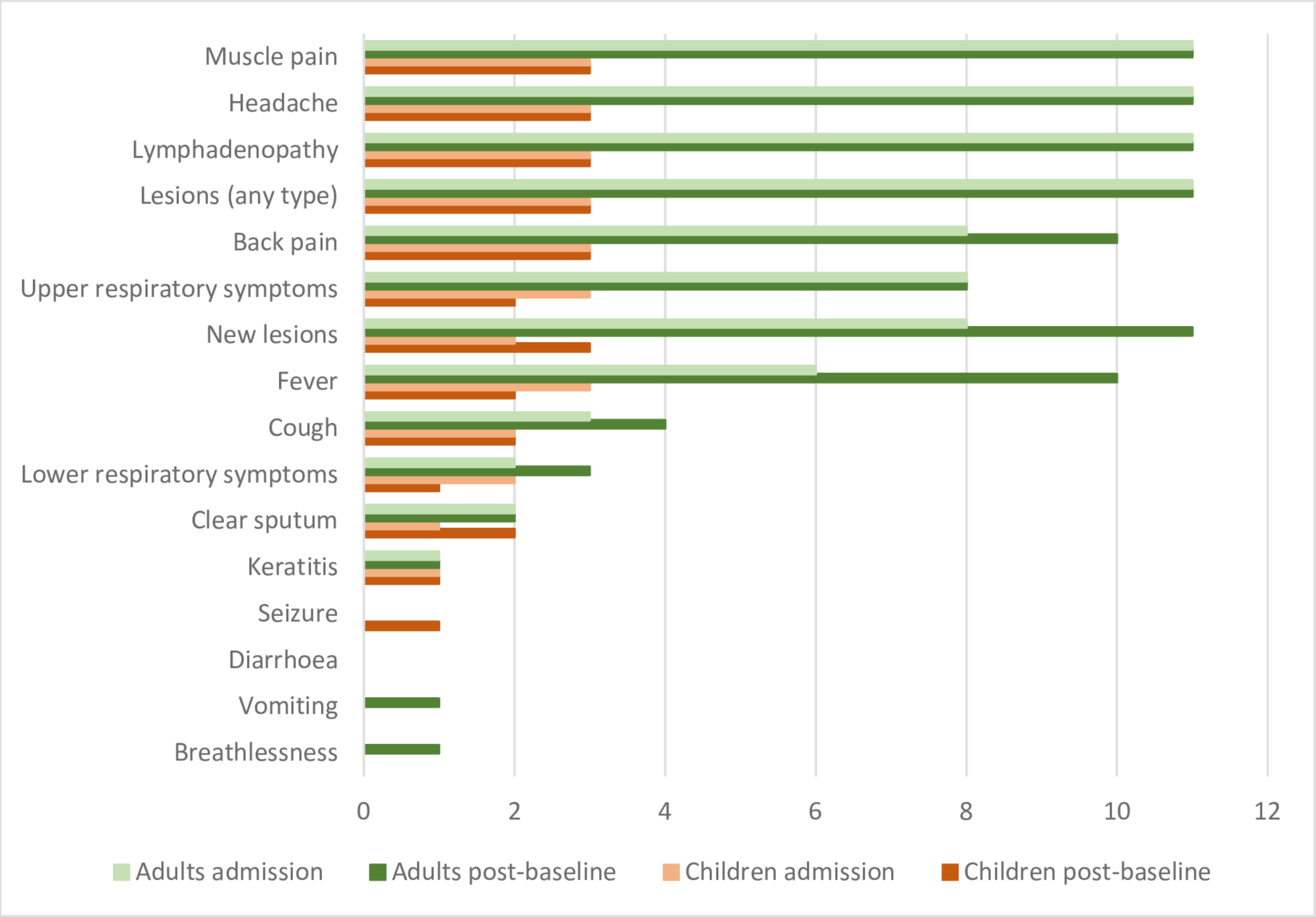

**Table S2 – Virological outcomes using G2R-G real-time PCR assay**

| | N patients tested | Results | CT values | | | |
|---|---|---|---|---|---|---|
| | | Positive n (%) | Mean | Std | Min | Max |
| **Baseline** | | | | | | |
| **Overall** | **14** | **14 (100%)** | | | | |
| Blood | 14 | 12 (86%) | 32 | 6 | 20 | 41 |
| Active lesion | 5 | 5 (100%) | 28 | 9 | 18 | 39 |
| Crust | 1 | 1 (100%) | | | | |
| **Day 4** | | | | | | |
| **Overall** | **14** | **7 (50%)** | | | | |
| Blood | 13 | 6 (46%) | 38 | 1 | 36 | 39 |
| Active lesion | 2 | 2 (100%) | 17 | 2 | 16 | 19 |
| Crust | 1 | 1 (100%) | | | | |
| **Day 8** | | | | | | |
| **Overall** | **10** | **1 (10%)** | | | | |
| Blood | 10 | 1 (10%) | | | | |
| Active lesion | 0 | 0 (0%) | | | | |
| Crust | 0 | 0 (0%) | | | | |
| **Day 14** | | | | | | |
| **Overall** | **8** | **1 (13%)** | | | | |
| Blood | 8 | 1 (13%) | | | | |
| Active lesion | 1 | 1 (100%) | | | | |
| Crust | 1 | 1 (100%) | | | | |
| **Day 21** | | | | | | |
| **Overall** | **2** | **1 (50%)** | | | | |
| Blood | 2 | 1 (50%) | | | | |
| Active lesion | 0 | 0 (0%) | | | | |
| Crust | 1 | 1 (100%) | | | | |
| **Final visit** | | | | | | |
| **Overall** | **13** | **1 (8%)** | | | | |
| Blood | 13 | 1 (8%) | | | | |
| Active lesion | 1 | 1 (100%) | | | | |
| Crust | 0 | 0 (0%) | | | | |

**Table S3 – Virological outcomes using C3L real-time PCR assay**

| | N patients tested | Result | CT values | | | |
|---|---|---|---|---|---|---|
| | | Positive n (%) | Mean | Std | Min | Max |
| **Baseline** | | | | | | |
| **Overall** | **14** | **12 (86%)** | | | | |
| Blood | 14 | 10 (71%) | 30 | 5 | 21 | 39 |
| Active lesion | 5 | 5 (100%) | 30 | 9 | 20 | 41 |
| Crust | 1 | 1 (100%) | | | | |
| **Day 4** | | | | | | |
| **Overall** | **14** | **5 (36%)** | | | | |
| Blood | 13 | 4 (31%) | 38 | 1 | 36 | 40 |
| Active lesion | 2 | 2 (100%) | 16 | 1 | 15 | 17 |
| Crust | 1 | 1 (100%) | | | | |
| **Day 8** | | | | | | |
| **Overall** | **10** | **1 (10%)** | | | | |
| Blood | 10 | 1 (10%) | | | | |
| Active lesion | 0 | 0 (0%) | | | | |
| Crust | 0 | 0 (0%) | | | | |
| **Day 14** | | | | | | |
| **Overall** | **8** | **1 (13%)** | | | | |
| Blood | 8 | 1 (13%) | | | | |
| Active lesion | 1 | 1 (100%) | | | | |
| Crust | 1 | 1 (100%) | | | | |
| **Day 21** | | | | | | |
| **Overall** | **2** | **1 (50%)** | | | | |
| Blood | 2 | 1 (50%) | | | | |
| Active lesion | 0 | 0 (0%) | | | | |
| Crust | 1 | 1 (100%) | | | | |
| **Final visit** | | | | | | |
| **Overall** | **13** | **1 (8%)** | | | | |
| Blood | 13 | 1 (8%) | | | | |
| Active lesion | 1 | 1 (100%) | | | | |
| Crust | 0 | 0 (0%) | | | | |

**Fig S4 – CT values of six patients from baseline to D4**

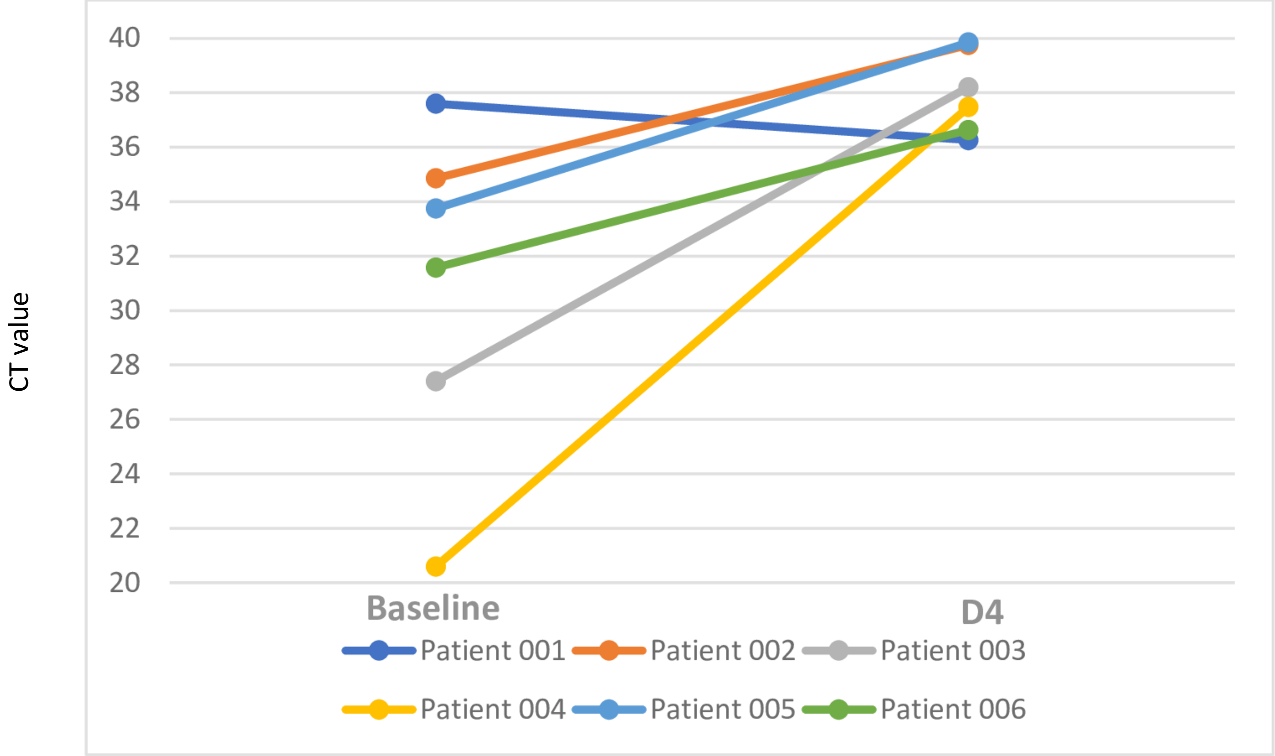
